# An alternative approach for bioanalytical assay development for wastewater-based epidemiology of SARS-CoV-2

**DOI:** 10.1101/2021.02.12.21251626

**Authors:** Boogaerts Tim, Lotte Jacobs, Naomi De Roeck, Siel Van den Bogaert, Bert Aertgeerts, Lies Lahousse, Alexander L.N. van Nuijs, Peter Delputte

## Abstract

Wastewater-based epidemiology could be applied to track down SARS-CoV-2 outbreaks at high spatio-temporal resolution and could potentially be used as an early-warning for emergence of SARS-CoV-2 circulation in the general population. Epidemiological surveillance of SARS-CoV-2 could play a role in monitoring the spread of the virus in the population and controlling possible outbreaks. However, sensitive sample preparation and detection methods are necessary to detect trace levels of SARS-CoV-2 RNA in influent wastewater (IWW).

Unlike predecessors, method development of a SARS-CoV-2 RNA concentration and detection procedure was performed with IWW samples with high viral SARS-CoV-2 loads (in combination with seeding IWW with a surrogate coronavirus). This is of importance since the SARS-CoV-2 genome in IWW might have already been subject to in-sewer degradation into smaller genome fragments or might be present in a different form (e.g. cell debris,…). Centricon Plus-70 (100 kDa) centrifugal filter devices resulted in the lowest and most reproducible Ct-values for SARS-CoV-2 RNA. Lowering pore sizes did not improve our limit of detection and quantification. Real-time polymerase chain reaction (qPCR) was employed for the amplification of the N1, N2, N3 and E_Sarbeco-gene.

This is one of the first studies to apply digital polymerase chain reaction (dPCR) for the detection of SARS-CoV-2 RNA in IWW. Interestingly, qPCR results were comparable with dPCR results suggesting that qPCR is a valid method. In this study, dPCR was also used as a proxy to assess the precision of qPCR. In this light, dPCR showed high variability at low concentration levels (10^0^ copies/µL), indicating that variability in bioanalytical assays for SARS-CoV-2 RNA might be substantial.

On average, the N2-gene showed high in-sample stability in IWW for 10 days of storage at 4 °C. Between-sample variability was substantial due to the low native concentrations in IWW. Additionally, the E-gene proved to be less stable compared to the N2-gene and showed higher variability. Freezing the IWW samples resulted in a 10-fold decay of loads of the N2- and E-gene in IWW.

Although WBE can already aid in filling some knowledge gaps in the epidemiological surveillance of SARS-CoV-2, future WBE studies should aim to further validate and standardize bioanalytical assays, especially with regards to methodological limitations.

**Highlights:**

- Development of an analytical procedure for detection of SARS-CoV-2 RNA in wastewater
- Extraction recovery was evaluated in influent wastewater
- Precision measured with dPCR used as a proxy for qPCR
- qPCR of the N2 gene fragment showed high in-sample stability of SARS-CoV-2 on average

## 1. Introduction

The Severe Acute Respiratory Syndrome Coronavirus 2 (SARS-CoV-2) is an enveloped non-segmented positive-sense RNA virus, which is associated with the pathogenesis of the coronavirus disease 2019 (COVID-19) in humans [1]. Due to the partly asymptomatic transmission and the high infectivity of this virus [2,3], it is crucial to have timely and accurate figures on the spread of SARS-CoV-2 in defined population groups for controlling possible viral outbreaks. Currently, the extent of SARS-CoV-2 circulation has been monitored by diagnostic testing of primarily symptomatic patients and contact tracing to also isolate asymptomatic patients [4,5]. However, a major limitation with these methods is that they depend on participation of individuals, even when they have no symptoms or only mild aspecific symptoms of COVID-19. Lack of recognition of symptoms or refusal to participate in detection or quarantine measures allows further spread of the virus in the general population. In Belgium, contact-tracing is primarily done manually through regional call centers and through the implementation of a smartphone application. Participation is heavily influenced by personal, social and public trust and requires additional efforts to connect with lower educated and vulnerable population groups [4]. Contact-tracing efforts could potentially be biased by reporting and concealment bias and requires from each individual to keep track of their contact list. Additionally, if contact-tracers fail to track down an individual’s contacts swiftly (for example due to the prolonged incubation period or time to perform diagnostic testing), it could have limited effect on the spread of this highly infectious virus [6].

Wastewater-based epidemiology (WBE) employs the analysis of influent wastewater (IWW) on human (metabolic) excretion products and has been used as an alternative approach to investigate the circulation and spread of infectious diseases at the population level (Figure 1) [7,8]. Infectious disease biomarkers (e.g. viral genomes) are released, pooled and transported in the wastewater system. The abundance of pathogens in IWW reflects the pattern and spread of infection at the population level and does not depend on participation at the individual level [9]. For this reason, WBE is an efficient alternative approach for the prevention of infectious disease outbreaks, to track down possible hotspots and to evaluate the effectiveness of large scale anticontagion interventions within different communities [10]. In order for WBE to monitor infectious diseases, the corresponding pathogen should preferentially be causing an enteric infection. Alternatively the pathogen or its genome should be excreted at sufficient levels in the faeces, urine or other excretions that end up in the wastewater [10–12]. In this light, several research papers indicated the potential of WBE in surveilling the transmission of the recently emerged SARS-CoV-2 at the population level [13–15].

**Figure 1.**
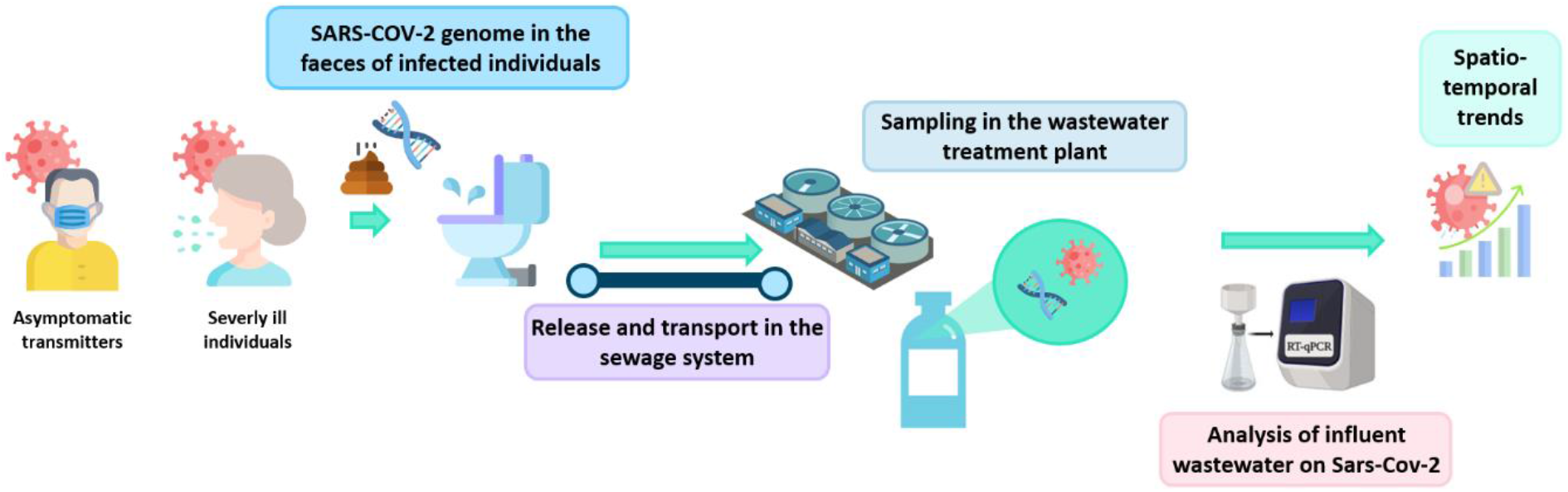
Schematic overview of sewage surveillance for determining SARS-CoV-2 circulation in the general population.

Thanks to some advantages of this epidemiologic approach, WBE could aid in filling some knowledge gaps. Asymptomatic individuals infected with SARS-CoV-2 also shed the virus via their stool in the sewers [11,16] enabling WBE to track down the extent of transmission of the virus in different catchments and to predict future disease outbreaks in communities. Additionally, WBE could potentially be used as an early-warning for emergence of SARS-CoV-2 in the general population [17–19]. WBE is especially useful to invigilate new outbreaks as it is highly inefficient to test thousands of individuals when detection rates are low. In this light, it is crucial to have sensitive and robust bioanalytical methods to quantitively measure SARS-CoV-2 in IWW.

The surveillance of wastewater on viral RNA loads of SARS-CoV-2 proposes some analytical challenges. These primarily include the low detection levels of the virus in IWW and the wastewater matrix potentially harboring a wide array of organic matter, humic acids and heavy metals that could interfere with the molecular methods of assaying viruses. Therefore, it is crucial to have reproducible concentration methods and sensitive instrumental techniques to accurately measure viruses in IWW [9]. At this moment, there has been a broad range of concentration methods for the extraction of viral loads of SARS-CoV-2 in IWW. Pre-existing concentration methods for SARS-CoV-2 RNA primarily include ultracentrifugation, ultrafiltration, charged filter membranes or PEG precipitation [20]. Nevertheless, most of these methods assess efficiency of recovery (RE) by spiking non-enveloped human enteric viruses to IWW [21–23]. Only a limited number of papers on method development and optimization investigate RE through spiking of enveloped viruses (e.g. coronavirus, the murine hepatitis virus and the bacteriophage pseudomonas virus phi6) [24]. However, even with enteric viruses, recoveries with concentrations methods are mostly determined by the virus and the matrix composition. In this light, coronaviruses (CoV) have quite distinctive structural and physical properties compared to other enteric viruses [25]. Furthermore, it is not exactly known in what complex the SARS-CoV-2 RNA is present in faeces (virus particles, cellular fragments…) and viral loads may be broken down in smaller fragments during in-sewer transport. Therefore such spiking experiments with viruses, although useful, are probably not fully representative for evaluation of stability and extraction of real IWW.

The aim of this study was to compare different bioanalytical procedures for the concentration of SARS-CoV-2 RNA in IWW. This bioanalytical assay was optimized with IWW originating from 8 Belgian wastewater treatment plants (WWTPs) with confirmed native levels of SARS-CoV-2 RNA, in combination with spiking with an animal Coronavirus. Additionally, in-sample stability at different storage conditions was further investigated. Finally, this study applied, as one of the first, digital polymerase chain reaction (dPCR) for assaying SARS-CoV-2 RNA in IWW, in a direct comparison with traditional qPCR.

## 2. Materials and methods

### 2.1. Reagents and materials

UltraPureTM DEPC-Treated Water (RNase and DNase free, molecular biology grade) was obtained from ThermoFisher Scientific (Waltham, US). Ethanol was obtained from Aventor (Radnor, US). CELLSTAR® serological pipettes (50 mL) and filter tips were obtained from BioScience and Greiner Bio-One International, respectively. The Eppendorf 5910R Centrifuge (Aarschot, BE) was used for sample centrifugation. Centricon Plus-70 Centrifugal filters (100 kDa and 30 kDa) and Amicon® Ultra Centrifugal filters (50 kDa and 10 kDa), Macrosep Advance Centrifugal devices with Omega Membrane (100 kDa and 30 kDa) and Vivaspin 20 ultrafiltration units (100 kDa and 50 kDa) were purchased from Millipore (Burlington, US), Pall (New York, US) and Sartorius (Göttingen, DE), respectively. PEG 8000 and sodium chloride were acquired from Promega (Madison, US).

The QIAamp Viral RNA minikit, the RNeasy plus minikit and the RNeasy Powermicrobiome kit were obtained from QIAGEN (Hilden, DE). Automated RNA extraction was done with the Maxwell® RSC Instrument with the PureFood GMO and Authentication kit, both from Promega (Madison, US). Before the molecular assay, samples were further purified from potential PCR-inhibitors with the 2x SensiFAST™ Probe No-ROX One-Step Kit from Bioline (Cincinnati, US). Real-time polymerase chain reaction (qPCR) was performed with the LightCycler® 96 instrument from Roche (Bazel, CH). SARS-CoV-2 and porcine coronavirus (PRCV) primers were obtained from Integrated DNA Technologies (IDT, Coralville, US) [26,27]. 96-well PCR pre-plates were acquired from Applied Biosystems (Thermo Fischer Scientific, Foster City, US). Table 1 summarizes the sequences for the primers and probes used for qPCR. Digital polymerase chain reaction (dPCR) was done with the QIAcuity One 5-Plex from QIAGEN (Hilden, DE). Samples were purified with the QiAcuity One-Step Viral RT-PCR kit and concentrations were 400 nM for primers and 200 nM for probes.

**Table 1.**
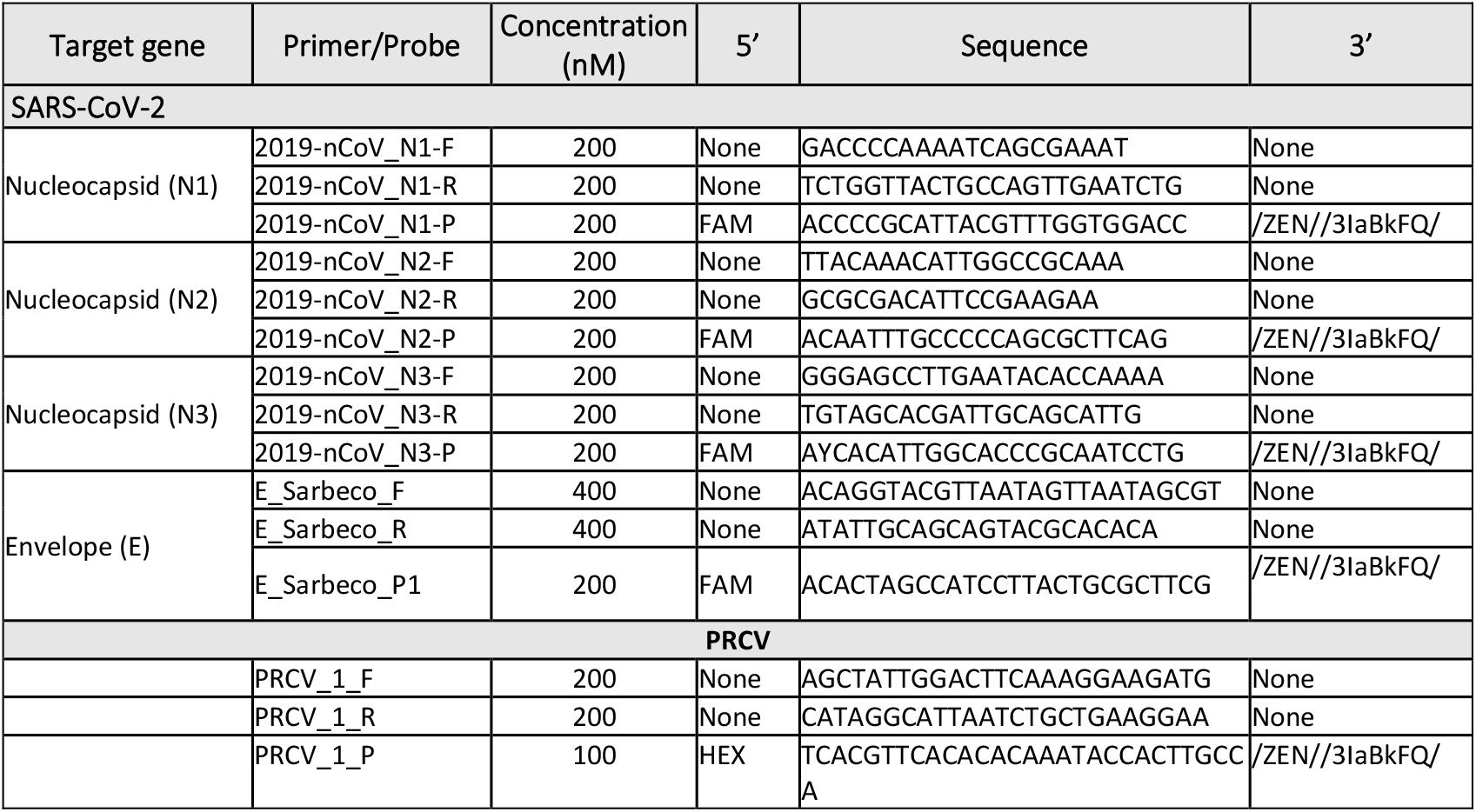
Real-time PCR primers and probes for the target virus and the whole process control

PRCV used as a whole process control was obtained from Ghent University (kind gift of Prof. H. Nauwynck, Merelbeke, Belgium). SARS-CoV-2 RNA used as a positive qPCR control was obtained from the Institute of Tropical Medicine Antwerp (kind gift of Prof. K. Ariën, ITG,BE). The EURM-019 reference standard for the construction of the calibration curve was obtained from the Joint Research Centre (JRC, European Commission). The Laboratory of Microbiology, Parasitology and Hygiene possesses the necessary permits for this research.

### 2.2. Sampling

Method development was done with IWW samples acquired from eight different Belgian WWTPs with population equivalents ranging between 25,000 to 200,000 inhabitants and sanitary wastewater (SAW) from a company that had a high number of positive COVID-19 cases (approximately 17% of the employers) [28]. Daily IWW samples were collected in the preamble (2^nd^ of August), peak (20^th^ of November) and tail (20^th^ of January) of the second wave of the COVID-19 pandemic. Locations are not further specified due to anonymity constraints, however, matrix compositions differ substantially between the locations of interest to demonstrate the robustness of the methodology. It should be noted that in-sewer degradation in the SAW is most likely less substantial compared to the IWW samples because of minor average residence times.

Daily 24-h composite IWW samples were collected time- or flow proportionally in order to obtain samples that were representative for an entire day. For time-proportional sampling, 10-minute intervals were applied to compile daily samples to accurately capture viral RNA loads over the 24-h period [29]. Average residence time was less than 12 hours in all locations. After sample collection, samples were transported immediately at 4 °C to the laboratory and analyzed within 24 hours.

### 2.3. Sample concentration

Virus concentration is necessary because of the low levels of SARS-CoV-2 RNA in wastewater. The analytical procedure needs to be sensitive enough to detect viral loads in the beginning or at the tail of the COVID-19 peak when only a limited number of SARS-CoV-2 infections are present in the catchment area. Several ultrafiltration methods (for protocols, see figures S1-S3) with different centrifugal devices with varying molecular weight cut-offs (MWCO) and loading volumes were tested for the concentration of viral RNA loads in IWW in order to obtain high extraction efficiencies. Additionally, PEG precipitation was also tested as an alternative for sample concentration. It should be noted that co-concentration of PCR inhibitors could also occur when using these concentration methods which could affect the assay’s sensitivity [23]. The composition of the IWW matrix is highly variable and contains a range of heavy metals, RNases and polysaccharides that could interfere with qPCR amplification.

It should be noted that RE was not determined by spiking the IWW samples pre- and post-extraction with an enveloped enteric control virus. In this study, an alternative approach was proposed in which IWW with substantial loads of SARS-CoV-2 RNA was tested with different sample concentration methods. The protocol with the lowest and most reproducible cycle threshold (Ct) levels for the amplification of the different SARS-CoV-2 genes was chosen for sample concentration. This approach was chosen since spiking IWW with enteric enveloped viruses may not be representative for RE of SARS-CoV-2 RNA in IWW due to different structural properties of these surrogate viruses or the in-sewer degradation of viral SARS-CoV-2 genome. Additionally, SARS-CoV-2 genome could also be present in a different form (e.g. cell debris) in the sewer system. However, during method development, IWW samples were spiked in parallel with PRCV to investigate whether the Ct-values for the amplification of the PRCV_1-gene were in line with the SARS-CoV-2 results when optimizing the different extraction protocols. Table 2 summarizes the design of experiment with the varying extraction protocols in order to obtain the most suitable sample concentration method of viral RNA loads in the wastewater matrix.

**Table 2.**
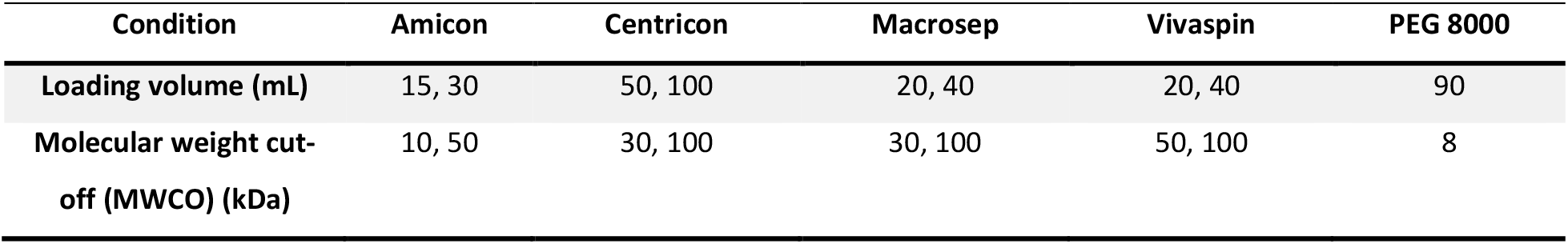
Optimizing a suitable sample concentration method

In the final protocol, samples were firstly centrifuged at 4625*g* for 30 minutes at 4 °C to remove solids and debris. The supernatans was transferred to a Centricon Plus-70 centrifugal filter for sample concentration. The sample was centrifuged for 15 minutes at 2500*g* at 4 °C in these centrifugal devices. Subsequently, the filter cup was centrifuged for an additional 2 minutes at 1000*g* at 4 °C to collect the sample concentrate. Finally, the sample concentrate was extracted and standardized at a volume of 1.5 mL.

### 2.4. RNA extraction

Similar to the sample concentration protocols, different commercially available manual RNA extraction kits were compared in order to obtain the lowest and most reproducible Ct-values for both SARS-CoV-2 and PRCV. Initially, a selection of IWW samples was processed with three manual RNA extraction kits (i.e. Viral RNA, RNeasy and Powermicrobiome). These RNA extraction kits need to be capable of isolating trace levels of SARS-CoV-2 RNA from IWW. The RE depends heavily on the composition of the RNA extraction kit. The number of washing steps to remove PCR inhibitors may vary between the kits and different elution solvents are used for the extraction of SARS-CoV-2 RNA from the concentrate. These commercialized kits have been proposed in other WBE applications for the extraction of SARS-CoV-2 in IWW [21].

For each RNA extraction kit, each sample concentration method was considered. The Ct-values obtained with the automated Maxwell PureFood GMO and Authentication RNA extraction kit were compared with the results from the manual RNA extraction kits to investigate whether it was possible to increase the throughput of the bioanalytical assay.

In the final method, extraction was performed with the automated Maxwell PureFood GMO and Authentication RNA extraction kit. 200 µL of the concentrate was added to 200 µL cetylrimethylammonium bromide buffer and 40 µL proteinase K and the total volume was incubated for 10 minutes at 56 °C. This mixture was transferred to the sample well together with 300 µL lysis buffer. The final elution volume with this RNA extraction kit was 50 µL.

### 2.5. Molecular methods for assaying SARS-CoV-2: qPCR and dPCR

All qPCR amplifications were performed in 20 µL reaction mixtures using a 2x SensiFAST™ Probe No-ROX One-Step kit for further purification from PCR-inhibitors after RNA extraction. Each reaction mixture contained 20% v/v of the extracted RNA. The final concentration of the primer and probes in the different qPCR mixtures was given in Table 1. A six-point calibration curve with a concentration between 10^5^ and 10^0^ copies/µL was constructed in ultrapure DEPC-treated water for quantification of the different genes of interest (Table 1) in IWW. The lower limit of quantification (LLOQ) was defined as the concentration in the lowest point of the calibration curve and was 10^0^ copies/µL for all genes. All qPCR reactions were performed in duplicate. For each qPCR run, two negative controls and a positive control were included. qPCR settings were as follows: 10 minutes for reverse transcription, 2 minutes for polymerase activation followed by 45 cycles of 5 seconds at 95 °C for denaturation and 30 seconds for annealing and extension.

In low-concentration IWW samples, qPCR could potentially be affected by sample inhibitors, poor amplification efficiency, less precision and the need for relative quantification, which might lead to the occurrence of false negative results. Therefore, dPCR was tested for the detection of SARS-CoV-2 RNA in IWW because of the higher precision, tolerance to PCR inhibitors and reproducibility compared to qPCR. dPCR could potentially be useful because of the low sensitivity to PCR inhibitors that are highly present in IWW and possibly co-concentrated during sample preparation. For this reason, this molecular assay could potentially be more sensitive for the detection of SARS-CoV-2 RNA in IWW. Another advantage of dPCR is that absolute quantification is performed (through Poisson calculations). The same standard curve was injected with dPCR for all genes of interest to investigate precision with dPCR at different concentration levels as a proxy for validation of the quantitative results. For each dPCR run a non-template control was included to determine the signal to noise (S/N) threshold.

### 2.6. Stability experiments

During transport in the sewer system, it is possible that the SARS-CoV-2 genome is broken down in smaller RNA fragments containing the SARS-CoV-2 genes. Therefore, in-sample stability was not investigated through standard addition with SARS-CoV-2 since stability of the corresponding gene fragments might not be comparable with the fragments found in IWW. This study only considered in-sample stability and in-sewer stability should be further investigated as medium to low stability could substantially influence the concentrations of SARS-CoV-2 RNA in the sewer system.

IWW samples from the eight different locations with substantial viral RNA loads were divided in multiple aliquots of 50 mL and stored at different temperatures (4 °C and −20 °C). These aliquots were subsequently analyzed at different time points for all genes of interest, as illustrated by Figure 2. Important to note is that all aliquots stored at −20 °C were only thawed once at the moment of analysis. The effect of multiple freeze-thaw cycles was not considered in this study. However, it has to be mentioned that multiple freeze-thaw cycles could lead to extensive breakdown of SARS-CoV-2 RNA by RNases present in IWW.

**Figure 2.**
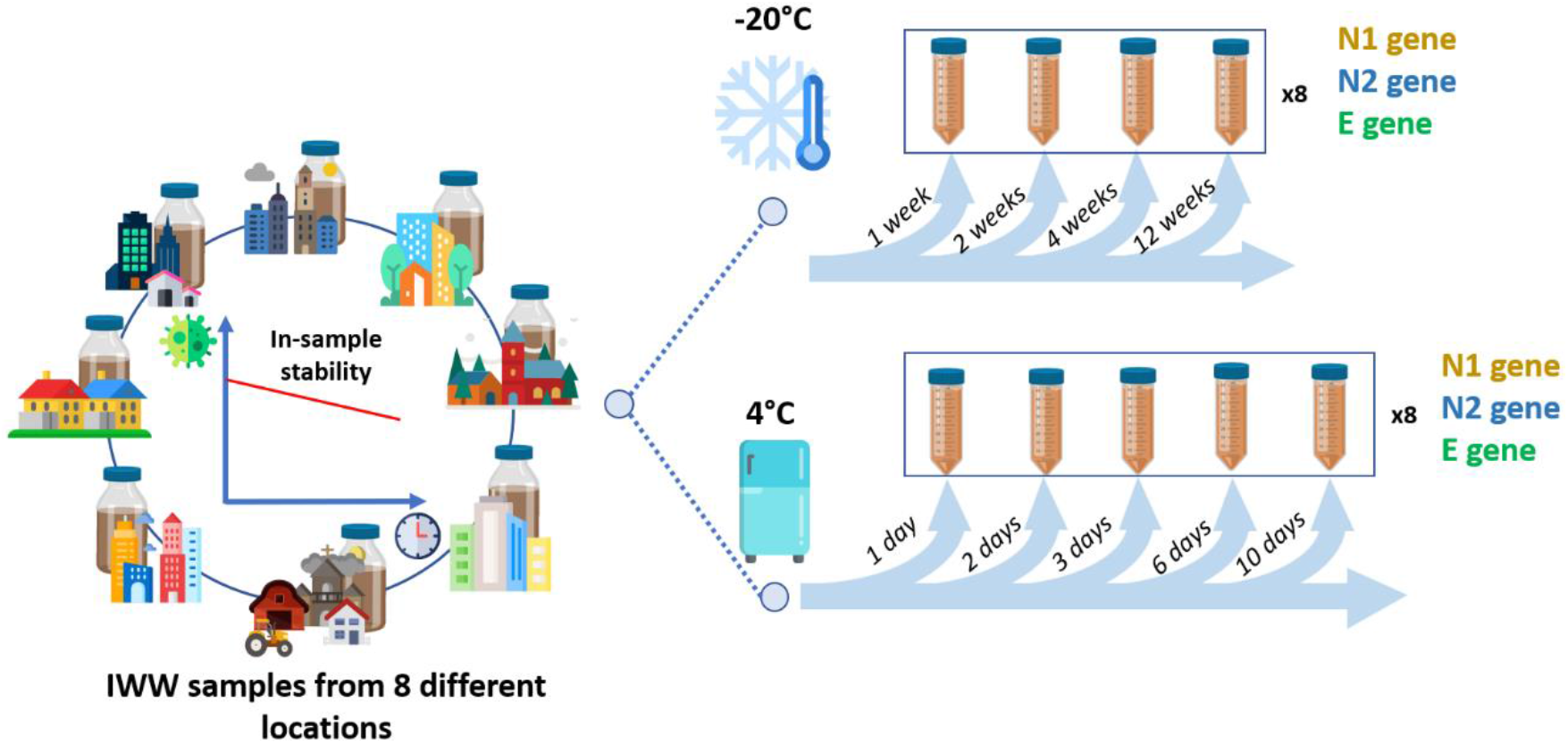
Schematic overview of the stability experiment.

For each IWW sample, viral loads for each gene were quantified at each time point and expressed as a relative percentage of the native concentration present in the corresponding IWW samples at the starting point of this stability study. The mean and relative standard deviation (%RSD) of all IWW samples were considered for each gene of interest.

## 3. Results & discussion

### 3.1. Sample concentration

Figures S1 summarizes the results of the different sample concentration methods for each RNA extraction kit. For this initial comparison, a fresh IWW sample (stored at 4 °C) from location 2 from the 2^nd^ of August 2020 was divided in different aliquots which were analyzed with different sample concentration methods for SARS-CoV-2 and seeded PRCV. At this time, the number of confirmed cases of COVID-19 was on the rise in this specific location, but was still considerably lower compared to the second wave of the pandemic. The Centricon Plus-70 centrifugal filters resulted in the highest yields of PRCV and SARS-CoV-2 for all genes of interest. While PEG precipitation resulted in detection levels of SARS-CoV-2 above the LLOQ for the E-gene, yields for the N1 and N2 gene were generally low or these genes remained undetected. This was in line with previous studies that reported poor recoveries of SARS-CoV-2 with PEG precipitation, possibly due to the co-concentration of PCR-inhibitors because of the small MWCO with this concentration method [21]. Ct-values measured with the other centrifugal methods were also low, as presented in Figure S4. However, yields of PRCV were considerably higher with the ultracentrifugation methods compared to the PEG precipitation. For this reason, PEG precipitation was excluded from further method optimization. The N3-gene resulted in poor detection levels with both RNA extraction kits and across all sample concentration methods. For this reason, further amplification of SARS-CoV-2 genes in IWW mainly focused on the N1-, N2- and E-gene.

The effect of different pore sizes and different loading volumes was tested with the different ultracentrifugation methods in order to obtain the highest and most reproducible RE for SARS-CoV-2. SAW from a company with high prevalence of SARS-CoV-2 infections was used for the optimization of the method, as illustrated by Figure 3A. Overall, increasing the sample volume resulted in the lowest Ct-values for the genes of interest with the different sample concentration methods. However, higher loading volumes (i.e. two times the maximum capacity of the filter) often resulted in blockage of the centrifugal filter membrane and, therefore, the IWW sample was only loaded once to prevent this. Blockage of the filter could also potentially lead to higher concentrations of PCR-inhibitors which could negatively influence the sensitivity with qPCR. The use of lower sample volumes also increases the throughput of the bioanalytical assay, since higher loading volumes require multiple centrifugation steps. Lower pore sizes did also not result in more sensitive detection of SARS-CoV-2; only the Amicon centrifugal filters with a MWCO of 10 kDa showed minor improvements. While sample concentration might be better with smaller pore sizes, decreasing the MWCO could also potentially lead to co-concentration of PCR-inhibitors. For this reason, MWCO ranging between 50 and 100 kDa were used for sample concentration. Concentration with the Centricon Plus-70 centrifugal filters resulted in the lowest Ct-values; the other centrifugal filters were comparable. Other studies also found acceptable RE of surrogate viruses or seeded SARS-CoV-2 in IWW with some of these ultracentrifugation methods. Loading volumes ranged between 50 and 500 mL in these studies. However, the variation of the RE with these sample concentration methods was quite substantial [21,23].

**Figure 3.**
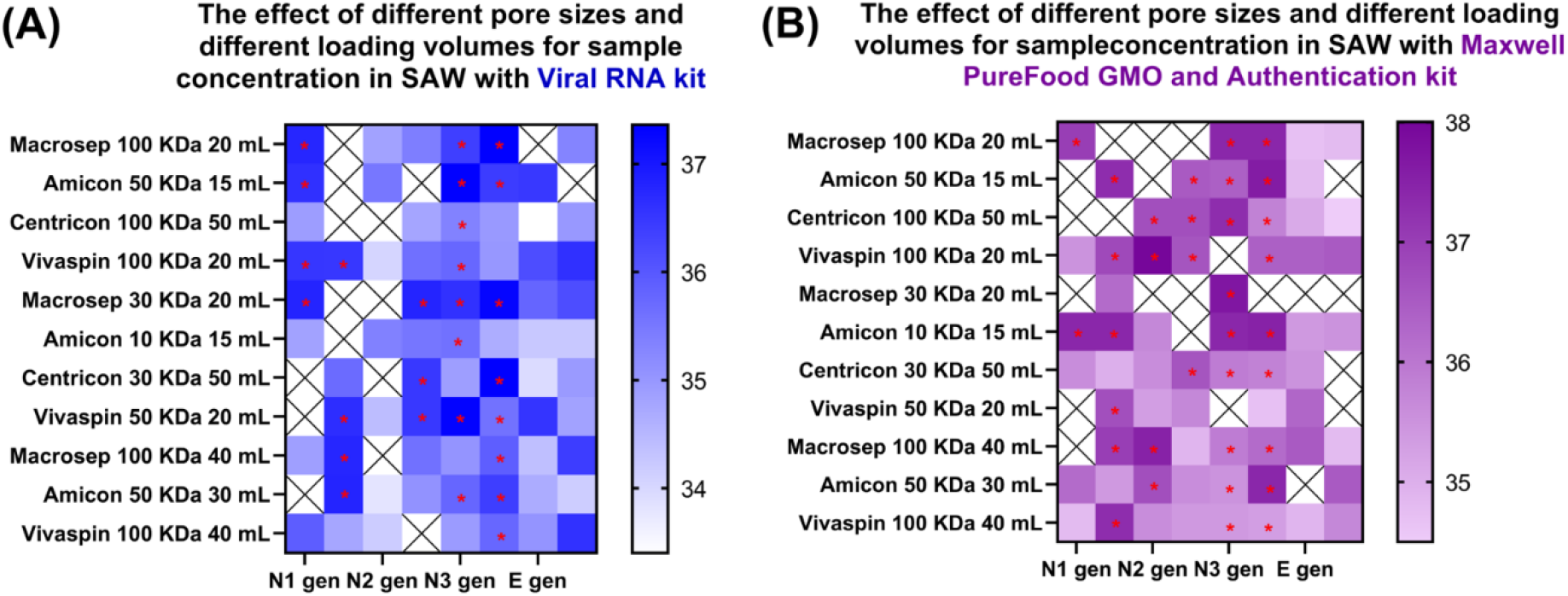
Optimization of pore sizes and loading volumes with the different sample concentration methods in SAW using (A) the Viral RNA extraction kit and (B) the Maxwell PureFood GMO and Authentication kit. The colour of each cell represents the Ct-value. Cells indicated with a red asterisk have higher Ct-values than the lowest point of the calibration curve and could therefore not be quantified. However, in these cells a positive signal was still detected. No signal was detected in cells with a black cross. Side-by-side cells for each location represent duplicate Ct-values.

Structural properties of the SARS-CoV-2 RNA found in the SAW might differ from the viral loads measured in IWW due to in-sewer degradation of viral RNA during sewage transport. Therefore, IWW samples (50 mL) collected from eight different Belgian WWTPs were processed with the Centricon (100 kDa) and the Macrosep (100 kDa) centrifugal filters to confirm the results. The effect of direct extraction was also considered because of the high incidence of SARS-CoV-2 infections at the time of sample collection. Figure 4 illustrates the Ct-values for the different SARS-CoV-2 genes under investigation for the different sample concentration methods. The use of no concentration step resulted in poor yields of SARS-CoV-2 RNA with Ct-values above the LLOQ only detected in a few locations, mainly for the N2-gene. This verifies the need for a concentration step to detect the low concentration levels of SARS-CoV-2 in IWW. The use of the Centricon centrifugal filters resulted in the lowest Ct-values for the N1- and E-gene. Results for the N2-gene were comparable between the Centricon and Macrosep centrifugal devices. In the final protocol, ultracentrifugation with Centricon filters was chosen for sample concentration.

**Figure 4.**
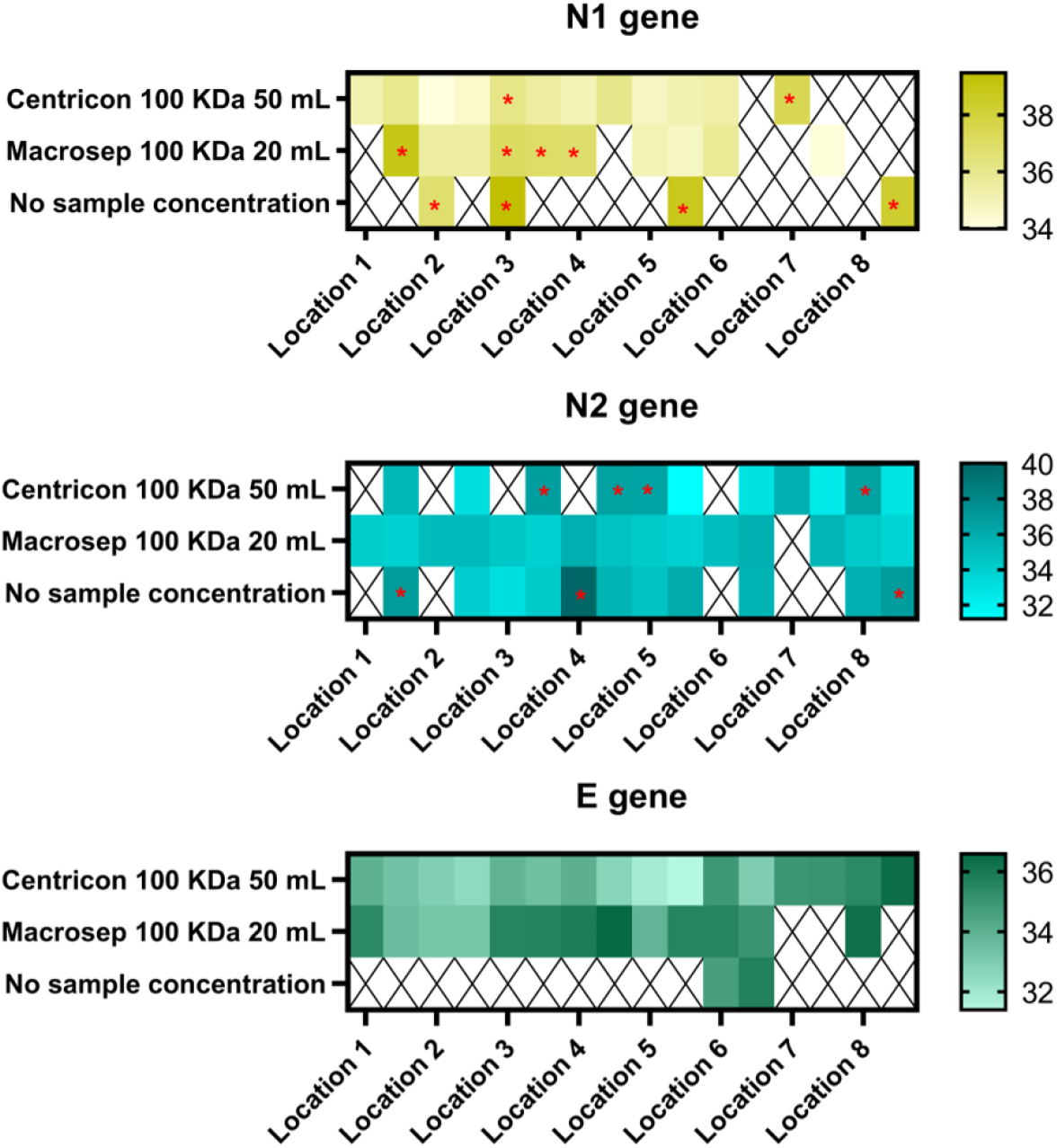
Comparison of sample concentration methods in IWW from 8 Belgian WWTPs. The colour of each cell represents the Ct-value. Cells indicated with a red asterisk have higher Ct-values than the lowest point of the calibration curve and could therefore not be quantified. However, in these cells a positive signal was still detected. No signal was detected in cells with a black cross. Side-by-side cells for each location represent duplicate Ct-values. The Maxwell PureFood GMO and Authentication kit was used for RNA extraction.

### 3.2. RNA extraction

Figure S4 compares the different RNA extraction protocols for both SARS-CoV-2 and PRCV. The use of the Powermicrobiome kit resulted in low detection levels of the different SARS-CoV-2 genes in IWW. Therefore, this RNA extraction method was excluded at an early stage. The Viral RNA and RNeasy extraction kit showed comparable results, with slightly higher detection levels observed with the Viral RNA extraction kit for both SARS-CoV-2 and PRCV. The Viral RNA extraction kit also recovered higher viral RNA loads in frozen IWW compared to the RNeasy extraction kit. However, in frozen IWW viral loads were almost always lower than the LLOQ, as further discussed in section 3.4.

Figure 3 provides a comparison between the manual Viral RNA extraction kit and the automated Maxwell PureFood GMO and Authentication kit. The use of both RNA extraction kits was comparable with slightly lower and more reproducible Ct-values among duplicates observed with the manual RNA extraction kit. However, automatization of the RNA extraction also significantly increases the throughput of the bioanalytical assay. For this reason, the Maxwell PureFood GMO and Authentication kit was used for RNA extraction in the final protocol.

### 3.3. Molecular methods for assaying SARS-CoV-2

#### 3.3.1. qPCR

A surrogate coronavirus (i.e. PRCV) was used as a whole process control. There was an expected repetitive drop in the Ct-value for the SARS-CoV-2 genes and PRCV_1-gene when amplifying a 10-fold dilution series proving the applicability of qPCR for the detection of the genes of interest. SARS-CoV-2 genes were not detected in the negative controls and the positive control tested always positive.

#### 3.3.2. dPCR

At this moment, investigation of SARS-CoV-2 genes with dPCR remains underexplored [30,31]. IWW samples from the tail of the second wave of the COVID-19 pandemic were analyzed with both qPCR and dPCR. dPCR could potentially be more sensitive in measuring viral RNA loads in IWW because of the higher tolerance to PCR inhibitors. Due to the high number of partitions (approximately 25,000), chances are low that PCR inhibitors and SARS-CoV-2 RNA are partitioned in the same reaction well. The positive partition rate was generally low in the IWW samples (<0.05%). At the low concentration levels observed in these samples, the 95% confidence interval (CI) with dPCR was also quite broad, as illustrated in Table 3. Concentrations measured with qPCR were the same order of magnitude as those with dPCR, with the exception of sample 3 and 4 for the E-gene (i.e. 10-fold higher with qPCR). Overall this indicates that sensitivity of both molecular assays is comparable. D’Aoust et al. also reported that dPCR did not result in superior detection of SARS-CoV-2 compared to qPCR [31]. In their study sensitivity was even lower compared to qPCR.

**Table 3.** Comparison of qPCR and dPCR results for all genes of interest for seven IWW samples collected in the tail of the second wave. CI = 95% confidence interval. Matrix composition (i.e. number of PCR inhibitors) was different between the different samples.

| Sample | Real-time qPCR |  |  | Digital PCR |  |  |  |  |  |
| --- | --- | --- | --- | --- | --- | --- | --- | --- | --- |
| | N1<br>(copies/μL) | N2<br>(copies/μL) | E<br>(copies/μL) | N1<br>(copies/μL) | CI<br>(%) | N2<br>(copies/μL) | CI<br>(%) | E<br>(copies/μL) | CI<br>(%) |
| 1 | 0.17 | 1.16 | n.d. | 0.70 | 95.7 | 0.54 | 109.1 | 0.27 | 168.6 |
| 2 | n.d. | n.d. | n.d. | 0.13 | 274.4 | n.d. | - | 0.26 | 168.6 |
| 3 | 0.46 | 1.15 | 10.17 | 1.33 | 64.9 | 1.00 | 79.0 | 0.13 | 274.4 |
| 4 | n.d. | 0.10 | 13.57 | n.d. | - | n.d. | - | 0.13 | 274.4 |
| 5 | 0.08 | 0.50 | n.d. | n.d. | - | n.d. | - | n.d. | - |
| 6 | 0.20 | n.d. | n.d. | 0.14 | 274.4 | n.d. | - | n.d. | - |
| 7 | 0.17 | 0.74 | 1.27 | 1.77 | 56.3 | 0.53 | 109.1 | 0.40 | 274.4 |

The same calibration curve was processed with both qPCR and dPCR. A major advantage of dPCR is that it allows absolute quantification and, thus, no standard curve is needed. The different calibration points were amplified with dPCR as a proxy to validate variability at different concentration levels and Ct-levels with qPCR. This is of importance because native concentrations of SARS-CoV-2 RNA in IWW are in the low copies/µL range. Important to note is that precision is generally higher with dPCR compared to qPCR because of the higher tolerance to PCR inhibitors and the use of absolute quantification. Therefore, precision observed with dPCR is used as a proxy for qPCR, but most likely the actual variability with qPCR will be higher due to methodological differences.

Figure 5 combines the result for the standard curve observed with qPCR with the results of dPCR. At low concentration levels (10^−1^ to 10^0^ copies/µL), the width of the CI tends to be rather broad. For the E-gene, no positive partitions were measured in the reaction well containing the 10^0^ copies/µL calibration point. The width of the CI for the N1- and N2-gene at this concentration level was 79.0% and 95.7% respectively (see also Table S1). This further evidences the high variability observed at Ct-values around the LLOQ with qPCR and could potentially explain why only one single well out of the side-by-side duplicates tested positive for SARS-CoV-2. Of course, the high variability observed in WBE applications for SARS-CoV-2 has further implications for the analysis of temporal trends in SARS-CoV-2 infections, especially in catchment areas with low prevalence of COVID-19. This uncertainty is further explored in section 4.

**Figure 5.**
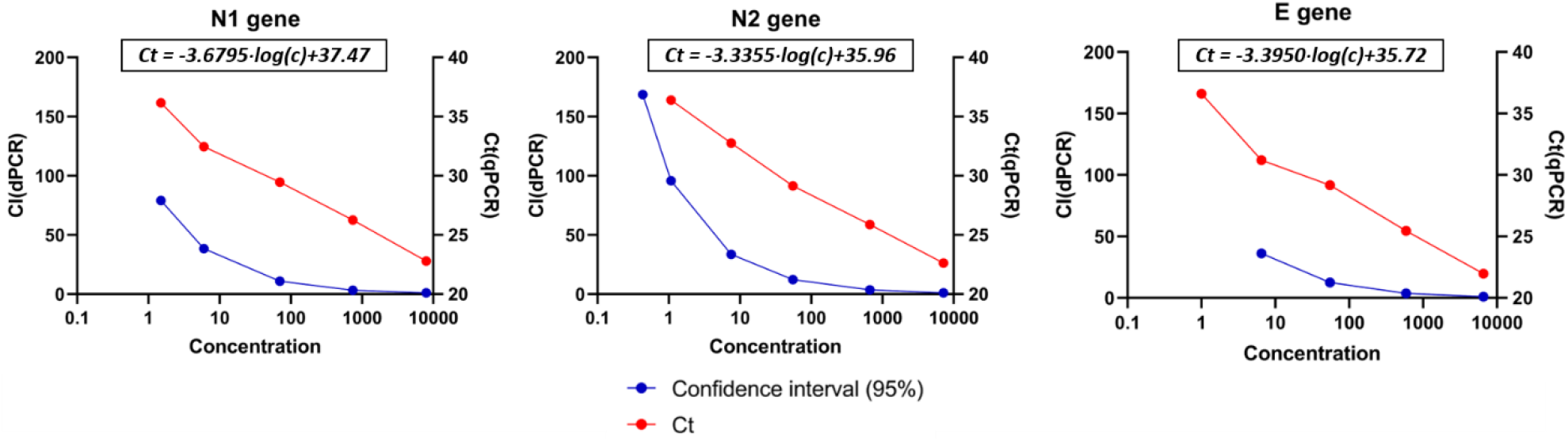
Evaluation of the precision of the standard curve with dPCR as a means to validate qPCR results. CI = confidence interval; Ct = cycle threshold. The left y-axis represents the precision of the 95% confidence interval after running the standard curve with dPCR. The right y-axis represents the Ct-values measured at the different concentration levels with qPCR.

### 3.4. Stability

In traditional WBE applications, stability is evaluated according to McCall et al. [32]. Generally, WBE biomarkers are classified as either high (<20% transformation), medium (20-60%), low (60-100%) or variable (i.e. different results found in WBE studies) stability over a pre-defined time period. However, the variability observed with native concentrations measured at the LLOQ levels (see section 3.3.2.) complicates this assessment.

The N2-gene and E-gene were detected in concentrations above the LLOQ in 100% and 87.5% of the IWW samples, respectively. The relative standard deviation (%RSD) was relatively high for both SARS-CoV-2 genes, as illustrated by Figure 6. Additionally, it appears that the variability for the E-gene tends to increase towards the end of the time period, with the highest %RSD values observed at day 10. On average, the N2-gene shows high in-sample stability for the entire sampling period, which is in line with the findings of others [17,33,34]. The E-gene showed medium to low in-sample stability during the time period, while others reported relatively high in-sample stability for this gene [33].

**Figure 6.**
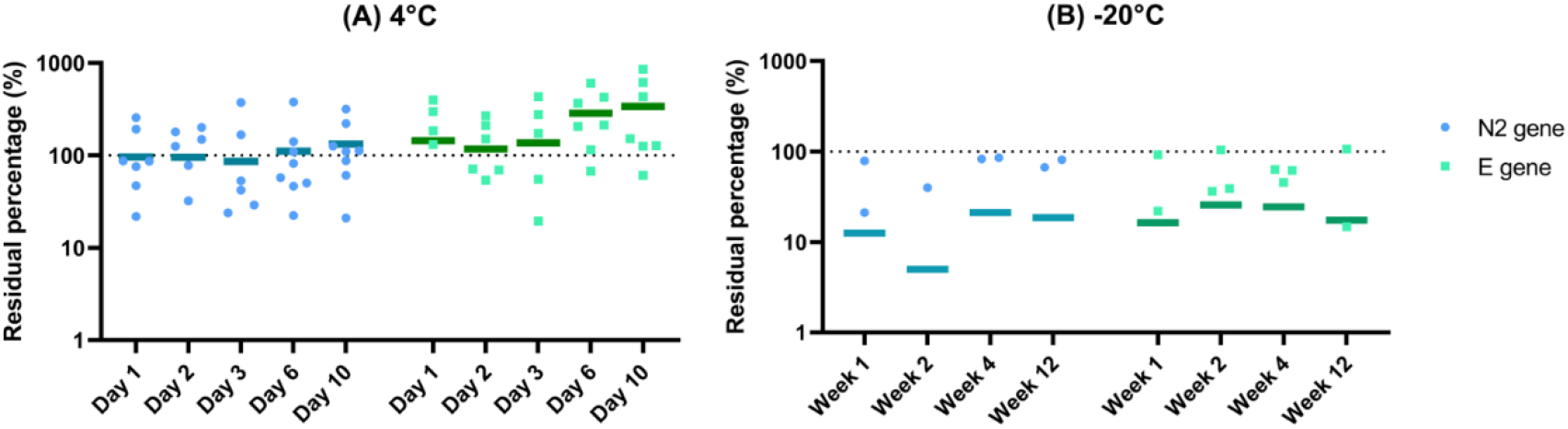
In-sample stability of the different SARS-CoV-2 genes at (A) 4 °C and (B) −20 °C. The horizontal lines represent the mean residual percentage for the eight samples. Detection of the N1-gene was generally low in the IWW samples (i.e. 10-1 copies/µL) with Ct-values below the LLOQ in most samples. Therefore, it was not possible to assess in-sample stability for the N1-gene.

Most of the stability studies use surrogate viruses to investigate the stability of SARS-CoV-2 in wastewater or often do not include all genes of interest [17,35,36]. Additionally, most of the available stability studies assess in-sample stability by seeding SARS-CoV-2 or another virus in IWW [33,34]. However, the genome of SARS-CoV-2 could potentially be degraded to smaller fragments during in-sewer transport. To our knowledge, only Hokajärvi et al. and Medema et al. investigated decay of native SARS-CoV-2 RNA loads in IWW [17,33] and variability reported by Hokajärvi was also substantial.

Freezing the samples drastically influences the in-stability of viral RNA loads of all genes of interest, with a 10-fold decrease in SARS-CoV-2 gene copies. This was also observed during method development (Figure S4) where detection levels were considerably higher in fresh IWW samples (kept at 4 °C) compared to frozen IWW samples.

During sample transport and storage, IWW samples should be kept at 4 °C to minimize in-sample degradation of viral RNA loads and IWW samples should be analyzed within three days after sample collection.

## 4. Uncertainties

At this moment, an ideal external control standard with the same properties as SARS-CoV-2 for quantification is missing [21]. In this study PRCV was used as a whole process control, but this surrogate might not entirely reflect the structural properties of the SARS-CoV-2 genome in IWW. In traditional WBE, isotope-labelled analogues are used for the quantification of chemical biomarkers. However, for biological applications, such controls are unavailable which could potentially lead to high variability with the current bioanalytical assays. Therefore, further methodological and molecular assay validation for SARS-CoV-2 RNA in IWW is required to enhance the accuracy and precision of WBE for SARS-CoV-2. This also further emphasizes the need for surrogate coronaviruses (e.g. PRCV) as whole process control to ensure overall quality of these bioanalytical assays. The presence of a whole process control is especially of importance because of the high variability in the composition of the matrix. The fraction of PCR inhibitors could vary within a single WWTP over time and is potentially very different between WWTPs. In this study, the %RSD at the low detection levels was still considerable, as indicated with dPCR. The high variability observed in the LLOQ range also addresses the need for replicates.

To our knowledge, no information is available on the in-sewer degradation of the SARS-CoV-2 genome. Fragmentation of the genome during in-sewer transport could potentially affect RE with the different concentration methods found in literature and lead to high variation between WWTPs due to different sewer structures. In the future, sequencing of the SARS-CoV-2 genome in IWW is required to identify the different SARS-CoV-2 strains in IWW.

In the final protocol, solids were removed during the pre-centrifugation of the IWW samples. However, adsorption of the SARS-CoV-2 genome to the pallet could affect RE. In this study, SARS-CoV-2 was detected to some extent in solids (data not shown), but the overall importance needs to be further explored.

## 5. Conclusions

The present study proposes an alternative approach to assess RE of SARS-CoV-2 genes in IWW with different ultracentrifugation protocols. Native concentration levels of the different SARS-CoV-2 genes measured in IWW from different Belgian WWTPs with the different sample concentration methods were used to optimize RE of SARS-CoV-2 RNA in IWW. The bioanalytical assay proved to be capable of measuring low concentrations of SARS-CoV-2 RNA present in the samples from different IWW sources. The present study is among the first to apply dPCR for the quantification of SARS-CoV-2 RNA in IWW and dPCR results were comparable with the qPCR results.

The variability observed with the sample concentration methods for SARS-CoV-2 remains substantial due to the lack of an ‘ideal’ external control standard with similar properties to SARS-CoV-2. At this moment, there is also a lot of uncertainty regarding the state of SARS-CoV-2 genome (fragments) in IWW due to potential in-sewer degradation. More research on variability of SARS-CoV-2 in IWW and potential transformation of SARS-CoV-2 RNA in the IWW is necessary to further investigate the applicability of WBE.

Although WBE can already aid in filling some critical knowledge gaps in the epidemiological surveillance of SARS-CoV-2, future research should aim to further validate and standardize bioanalytical assays, especially with regards to methodological uncertainties. This is especially of importance when the number of WBE applications on data triangulation with other epidemiological information sources outpace the number of WBE studies that investigate intrinsic methodological uncertainties.

## Data Availability

The data that support the findings of this study are given in the manuscript and supplementary information.

## Author contributions

**Tim Boogaerts** – Conceptualization, data curation, formal analysis, investigation, methodology, visualization, roles/writing – original draft, writing-review and editing

**Naomi De Roeck** – Conceptualization, Data curation, formal analysis, investigation, methodology, writing-review and editing

**Lotte Jacobs**-Data curation, formal analysis, investigation, methodology, writing-review and editing

**Siel Van Den Bogaert** – Data curation, writing – review and editing

**Bert Aertgeerts** – Writing – review and editing

**Lies Lahousse** – Writing – review and editing

**Alexander van Nuijs** – Conceptualization, Data curation, formal analysis, investigation, methodology, writing-review and editing, supervision, funding acquisition

**Peter Delputte** – Conceptualization, Data curation, formal analysis, investigation, methodology, writing-review and editing, supervision, funding acquisition

## Acknowledgements

The funding of this project was obtained from the Research Council (BOF) of the University of Antwerp [project number: FFB200184] and the Agency of Care and Health [project number: GE0-1GPFZJA-WT]. Additionally, we thank the personnel of Aquafin for their support in collection of the IWW samples.

## References

1. Huang C, Wang Y, Li X, et al. Clinical features of patients infected with 2019 novel coronavirus in Wuhan, China. Lancet. 2020;395(10223):497–506. doi:https://doi.org/10.1016/S0140-6736(20)30183-5

2. Chen N, Zhou M, Dong X, et al. Epidemiological and clinical characteristics of 99 cases of 2019 novel coronavirus pneumonia in Wuhan, China: a descriptive study. Lancet. 2020;395(10223):507–513. doi:https://doi.org/10.1016/S0140-6736(20)30211-7

3. Gao QY, Chen YX, Fang JY. 2019 Novel coronavirus infection and gastrointestinal tract. J Dig Dis. 2020;21(3):125–126. doi:https://doi.org/10.1111/1751-2980.12851

4. Vandamme A-M, Nguyen T. Belgium - concerns about coronavirus contact-tracing apps. Nature. 2020;581(7809):384. doi:10.1038/d41586-020-01552-w

5. Peccia J, Zulli A, Brackney DE, et al. Measurement of SARS-CoV-2 RNA in wastewater tracks community infection dynamics. Nat Biotechnol. 2020;38(10):1164–1167. doi:10.1038/s41587-020-0684-z

6. He X, Lau EHY, Wu P, et al. Temporal dynamics in viral shedding and transmissibility of COVID-19. Nat Med. 2020;26(5):672–675. doi:10.1038/s41591-020-0869-5

7. Mao K, Zhang K, Du W, Ali W, Feng X, Zhang H. The potential of wastewater-based epidemiology as surveillance and early warning of infectious disease outbreaks. Curr Opin Environ Sci Heal. 2020;17:1–7. doi:10.1016/j.coesh.2020.04.006

8. Sims N, Kasprzyk-Hordern B. Future perspectives of wastewater-based epidemiology: Monitoring infectious disease spread and resistance to the community level. Environ Int. 2020;139:105689. doi:10.1016/j.envint.2020.105689

9. Corpuz MVA, Buonerba A, Vigliotta G, et al. Viruses in wastewater: occurrence, abundance and detection methods. Sci Total Environ. 2020;745:140910. doi:https://doi.org/10.1016/j.scitotenv.2020.140910

10. Hamouda M, Mustafa F, Maraqa M, Rizvi T, Aly Hassan A. Wastewater surveillance for SARS-CoV-2: Lessons learnt from recent studies to define future applications. Sci Total Environ. 2021;759:143493. doi:10.1016/j.scitotenv.2020.143493

11. Zhang J, Wang S, Xue Y. Fecal specimen diagnosis 2019 novel coronavirus–infected pneumonia. J Med Virol. 2020;92(6):680–682. doi:https://doi.org/10.1002/jmv.25742

12. Pan Y, Zhang D, Yang P, Poon LLM, Wang Q. Viral load of SARS-CoV-2 in clinical samples. Lancet Infect Dis. 2020;20(4):411–412. doi:https://doi.org/10.1016/S1473-3099(20)30113-4

13. Wu F, Zhang J, Xiao A, et al. SARS-CoV-2 Titers in Wastewater Are Higher than Expected from Clinically Confirmed Cases. mSystems. 2020;5(4). doi:10.1128/mSystems.00614-20

14. La Rosa G, Iaconelli M, Mancini P, et al. First detection of SARS-CoV-2 in untreated wastewaters in Italy. Sci Total Environ. 2020;736:139652. doi:10.1016/j.scitotenv.2020.139652

15. Ahmed W, Angel N, Edson J, et al. First confirmed detection of SARS-CoV-2 in untreated wastewater in Australia: A proof of concept for the wastewater surveillance of COVID-19 in the community. Sci Total Environ. 2020;728:138764. doi:10.1016/j.scitotenv.2020.138764

16. Lescure F-X, Bouadma L, Nguyen D, et al. Clinical and virological data of the first cases of COVID-19 in Europe: a case series. Lancet Infect Dis. 2020;20(6):697–706. doi:https://doi.org/10.1016/S1473-3099(20)30200-0

17. Medema G, Been F, Heijnen L, Petterson S. Implementation of environmental surveillance for SARS-CoV-2 virus to support public health decisions: Opportunities and challenges. Curr Opin Environ Sci Heal. 2020;17:49–71. doi:https://doi.org/10.1016/j.coesh.2020.09.006

18. Orive G, Lertxundi U, Barcelo D. Early SARS-CoV-2 outbreak detection by sewage-based epidemiology. Sci Total Environ. 2020;732:139298. doi:10.1016/j.scitotenv.2020.139298

19. Ahmed W, Tscharke B, Bertsch PM, et al. SARS-CoV-2 RNA monitoring in wastewater as a potential early warning system for COVID-19 transmission in the community: A temporal case study. Sci Total Environ. 2021;761:144216. doi:10.1016/j.scitotenv.2020.144216

20. Kitajima M, Ahmed W, Bibby K, et al. SARS-CoV-2 in wastewater: State of the knowledge and research needs. Sci Total Environ. 2020;739:139076. doi:https://doi.org/10.1016/j.scitotenv.2020.139076

21. Alygizakis N, Markou AN, Rousis NI, et al. Analytical methodologies for the detection of SARS-CoV-2 in wastewater: Protocols and future perspectives. Trends Analyt Chem. 2021;134:116125. doi:10.1016/j.trac.2020.116125

22. Jafferali MH, Khatami K, Atasoy M, Birgersson M, Williams C, Cetecioglu Z. Benchmarking virus concentration methods for quantification of SARS-CoV-2 in raw wastewater. Sci Total Environ. 2021;755(Pt 1):142939. doi:10.1016/j.scitotenv.2020.142939

23. Ahmed W, Bivins A, Bertsch PM, et al. Surveillance of SARS-CoV-2 RNA in wastewater: Methods optimisation and quality control are crucial for generating reliable public health information. Curr Opin Environ Sci Heal. Published online September 2020. doi:10.1016/j.coesh.2020.09.003

24. Ye Y, Ellenberg RM, Graham KE, Wigginton KR. Survivability, Partitioning, and Recovery of Enveloped Viruses in Untreated Municipal Wastewater. Environ Sci Technol. 2016;50(10):5077–5085. doi:10.1021/acs.est.6b00876

25. Haramoto E, Kitajima M, Hata A, et al. A review on recent progress in the detection methods and prevalence of human enteric viruses in water. Water Res. 2018;135:168–186. doi:https://doi.org/10.1016/j.watres.2018.02.004

26. Centers for Disease Control and Prevention. Research Use Only 2019-Novel Coronavirus (2019-nCoV) Real-time RT-PCR Primers and Probes. Published 2020. https://www.cdc.gov/coronavirus/2019-ncov/lab/rt-pcr-panel-primer-probes.html

27. Corman V, Bleicker T, Brünick S, Drosten C. Diagnostic Detection of 2019-NCoV by Real-Time RT-PCR.; 2020. https://www.who.int/docs/default-source/coronaviruse/protocol-v2-1.pdf

28. Outters A. All employers of Westvlees tested: a total of 94 infections. Vrt News. https://www.vrt.be/vrtnws/nl/2020/08/14/alle-werknemers-van-westvlees-getest-in-totaal-94-mensen-besmet/. xPublished 2020.

29. Ort C, Lawrence MG, Reungoat J, Mueller JF. Sampling for PPCPs in Wastewater Systems: Comparison of Different Sampling Modes and Optimization Strategies. Environ Sci Technol. 2010;44(16):6289–6296. doi:10.1021/es100778d

30. Pecson BM, Darby E, Haas CN, et al. Reproducibility and sensitivity of 36 methods to quantify the SARS-CoV-2 genetic signal in raw wastewater: findings from an interlaboratory methods evaluation in the U.S. Environ Sci Water Res Technol. Published online 2021. doi:10.1039/D0EW00946F

31. D’Aoust PM, Mercier É, Montpetit D, et al. Quantitative analysis of SARS-CoV-2 RNA from wastewater solids in communities with low COVID-19 incidence and prevalence. medRxiv. Published online January 1, 2020:2020.08.11.20173062. doi:10.1101/2020.08.11.20173062

32. McCall A-K, Bade R, Kinyua J, et al. Critical review on the stability of illicit drugs in sewers and wastewater samples. Water Res. 2016;88:933–947. doi:10.1016/j.watres.2015.10.040

33. Hokajärvi A-M, Rytkönen A, Tiwari A, et al. The detection and stability of the SARS-CoV-2 RNA biomarkers in wastewater influent in Helsinki, Finland. Sci Total Environ. 2021;770:145274. doi:10.1016/j.scitotenv.2021.145274

34. Ahmed W, Bertsch PM, Bibby K, et al. Decay of SARS-CoV-2 and surrogate murine hepatitis virus RNA in untreated wastewater to inform application in wastewater-based epidemiology. Environ Res. 2020;191:110092. doi:https://doi.org/10.1016/j.envres.2020.110092

35. Casanova L, Rutala WA, Weber DJ, Sobsey MD. Survival of surrogate coronaviruses in water. Water Res. 2009;43(7):1893–1898. doi:https://doi.org/10.1016/j.watres.2009.02.002

36. Gundy PM, Gerba CP, Pepper IL. Survival of Coronaviruses in Water and Wastewater. Food Environ Virol. 2009;1(1):10. doi:10.1007/s12560-008-9001-6

