## Supplementary Information for "An alternative approach for bioanalytical assay development for wastewater-based epidemiology of SARS-CoV-2"

### S.1. Sample preparation

*S.1.1. Standard operating procedures for the different sample concentration methods*

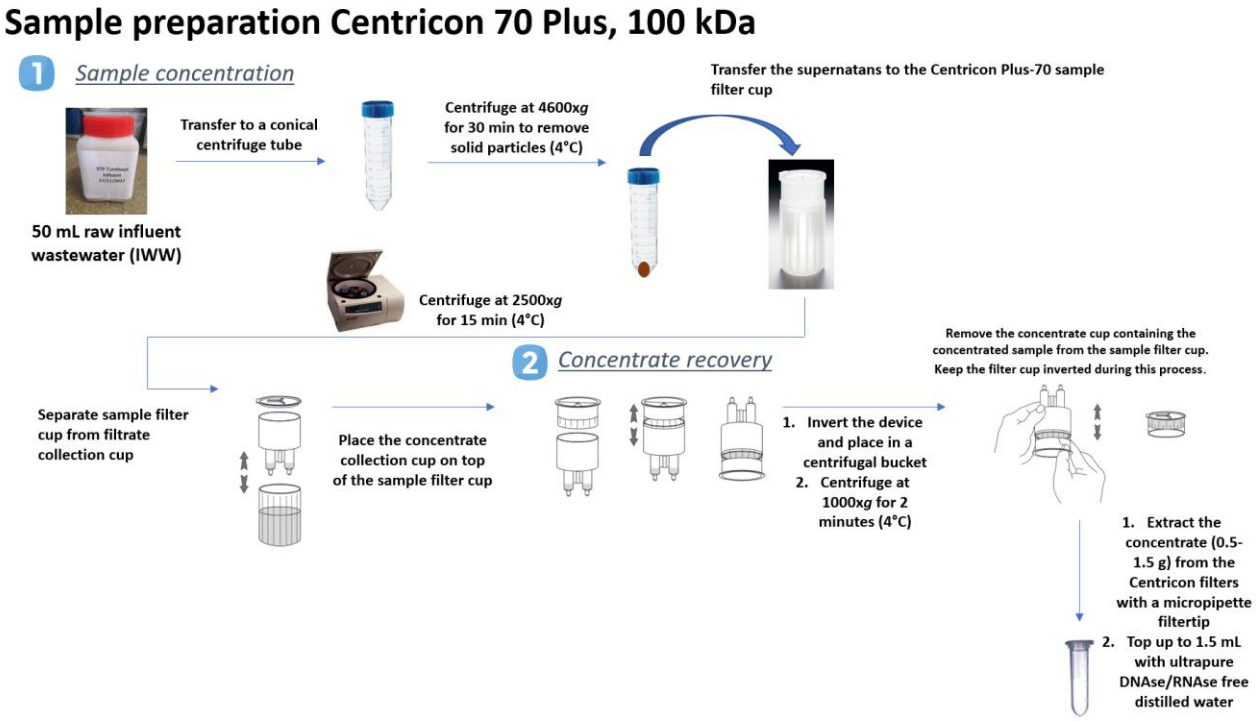


*Figure S 1 SOP Centricon Plus-70 centrifugal devices*

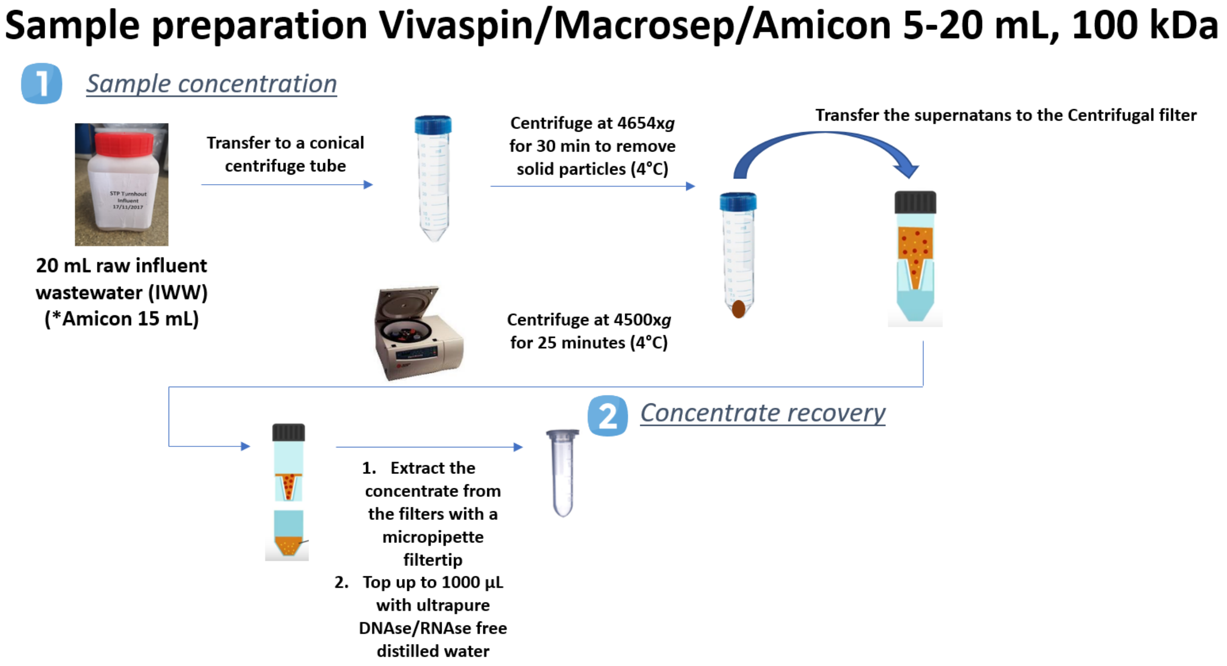


*Figure S 2 SOP centrifugal filter units (Vivaspin/Macrosep/Amicon)*

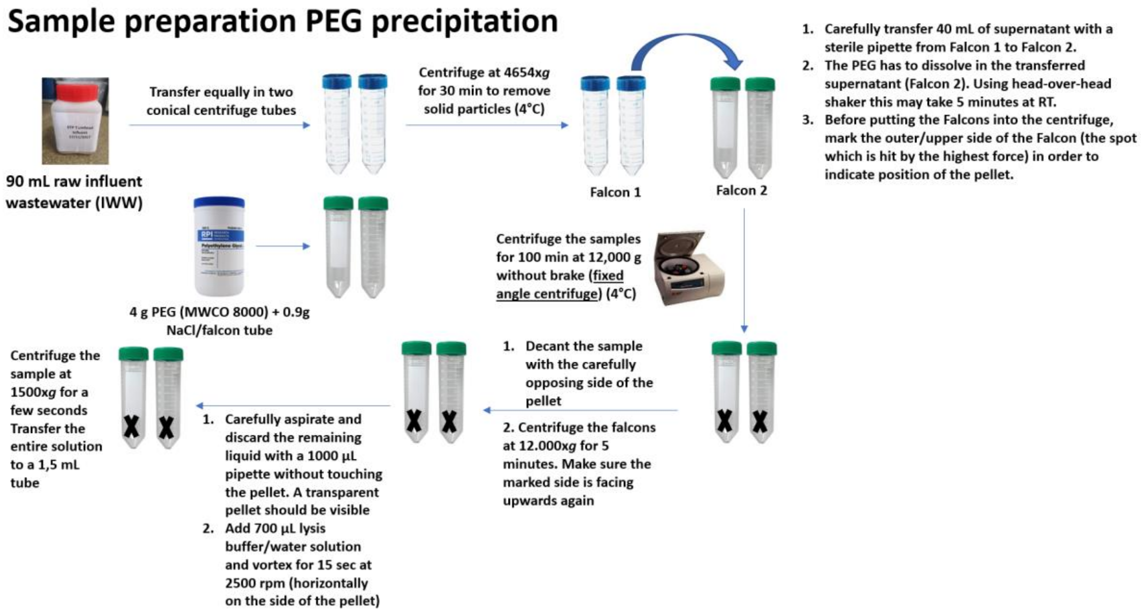


***Figure S 3 SOP PEG precipitation***

### *S.1.2. Comparison of different sample preparation methods*

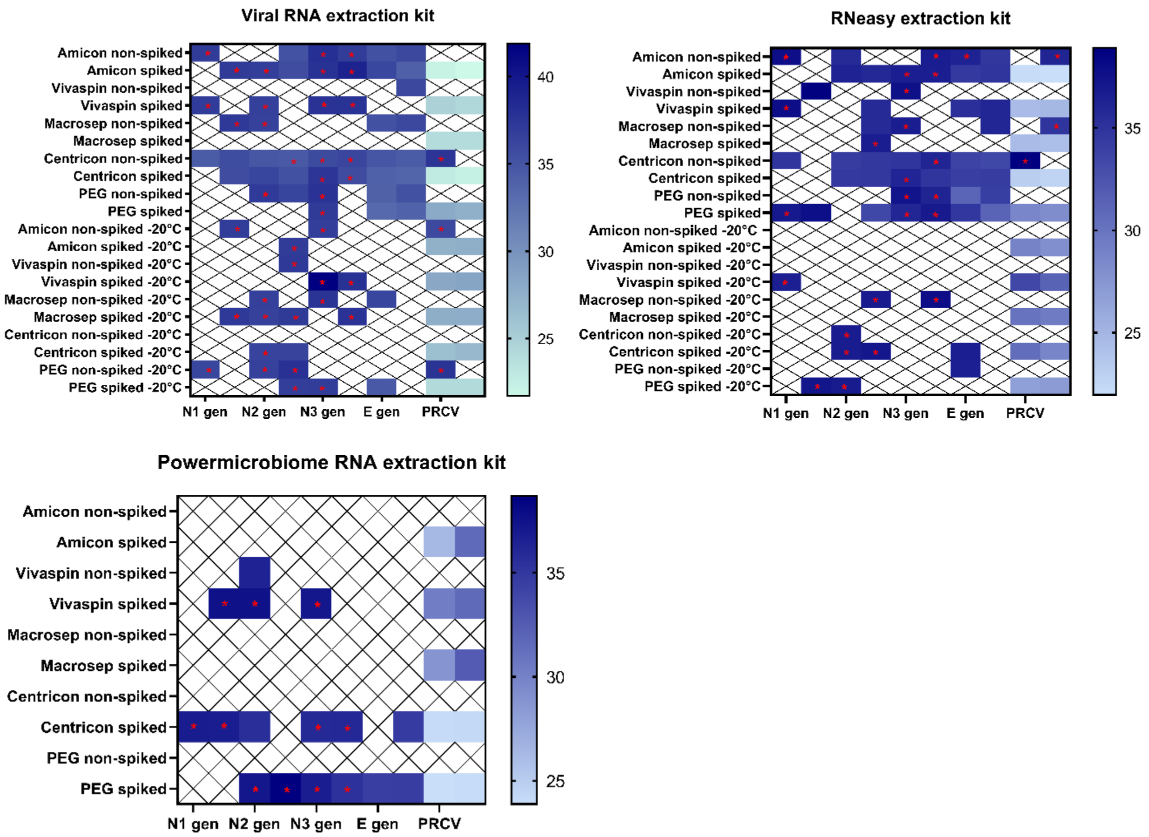


***Figure S 4 Comparison of different sample concentration methods and RNA extraction kits with both frozen (-20°C) and fresh (4°C) IWW samples.*** *The same sample was aliquoted in different fractions that were analyzed with the different sample concentration methods and RNA extraction kits.*

### S. 1.2. Instrumental analysis

***Table S 1 Assessment of precision at different concentration levels with dPCR***

| Calibration point | Target gene | Concentration (copies/µL) | Confidence interval (95%) | Number of valid partitions | Positive partition rate |
|---|---|---|---|---|---|
| | N1 | 7967 | 1.0% | 24882 | 76.6% |
| $10^4$ | N2 | 7212.5 | 1.1% | 25426 | 73.1% |
| | E | 6565.5 | 1.1% | 25173 | 69.1% |
| | N1 | 741 | 3.3% | 25459 | 12.6% |
| $10^3$ | N2 | 665.5 | 3.5% | 25469 | 11.4% |
| | E | 591.5 | 3.7% | 25451 | 10.2% |
| | N1 | 70 | 10.9% | 25453 | 1.2% |
| $10^2$ | N2 | 55.5 | 12.2% | 25428 | 1.0% |
| | E | 55 | 12.6% | 24116 | 1.0% |
| | N1 | 6 | 38.4% | 24817 | 0.10% |
| $10^1$ | N2 | 7.5 | 33.6% | 25460 | 0.13% |
| | E | 6.5 | 36.3% | 25451 | 0.11% |
| | N1 | 1.505 | 79.0% | 25468 | 0.027% |
| $10^0$ | N2 | 1.075 | 95.7% | 25460 | 0.020% |
| | E | 0 | - | 25439 | 0% |
| | N1 | 0 | - | 24753 | 0% |
| $10^{-1}$ | N2 | 0.43 | 168,6% | 25457 | 0.0079% |
| | E | 0 | - | 25446 | 0% |
